# A Pilot Meta-research on Evolving Evidence Behind Genetic Variant (Re)Classification

**DOI:** 10.1101/2025.04.07.25325116

**Authors:** Haotian Ma, Zihan Xu, Wendy Chung, Chunhua Weng, Yifan Peng

**Author notes:** These authors contributed equally to this work (Haotian Ma, Zihan Xu).

## Abstract

Variant classification and reclassification are fundamental to advancing precision medicine. This study focuses on the reclassifications of variants of uncertain significance (VUS) in *BRCA1* and *BRCA2* genes. By analyzing 162 unique cited publications supporting VUS reclassifications, we examined the accuracy, completeness, and currency of citations to these publications. Our findings reveal missing or inadequate evidence for reclassifications, as well as temporally misaligned citations and ClinVar submissions. Furthermore, we observed patterns in the cited studies, including the use of classification recommendations, genetic mechanisms, computational tools, and diverse population studies. This study underscores the need for stronger evidence supporting reclassifications and greater inclusion of diverse populations to optimize genomic variant reclassification and clinical decision-making.

## 1. Introduction

The classification of genomic variants as pathogenic, likely pathogenic, benign, likely benign, or variant of uncertain significance (VUS) is important in transforming genetic insights into clinical decision-making in the care of cancer and other medical conditions. For example, identifying a variant as pathogenic in genes such as *BRCA1* or *BRCA2* can influence breast cancer prevention and treatment, including risk-reducing surgeries, enhanced surveillance, or targeted treatments such as Poly Adenosine Diphosphate-Ribose Polymerase (PARP) inhibitors. In contrast, confirming variants as benign helps avoid unnecessary, intrusive, or risky interventions. Compared to variant classification, variant reclassification is particularly critical to resolving the ambiguity associated with VUS. As genomic research and technologies advance, variants initially classified as VUS may later be reclassified as pathogenic or benign, significantly impacting clinical management.

While there is a growing body of literature on variant reclassifications, few studies have systematically examined them to understand the variation among clinical sub-groups, methodology, classification guidelines, publication date, and other factors. This meta-research study aims to examine the published literature supporting variant reclassification. To achieve this, we identified variants whose classification has changed over time. We focus on two genes with significant clinical impact and a large amount of available data, BRCA1 and BRCA2. These two genes are essential to understanding breast cancer risk. In addition, BRCA1/2 variants are some of the most researched genes that set the basis for this study [1, 2]. Next, we assessed the classifications of variants in these two genes in ClinVar [3] and extracted the publications cited to support reclassifications. We analyzed the metadata of these studies, including study population and bioinformatics tools in use, as well as timelines of publications.

We identified two areas needing improvement for variant classification reporting. First, the clarity and relevance of evidence presented in ClinVar’s cited articles, and the quality were sometimes limited. Even when specific variants were mentioned, the classification was not always explicitly stated. Second, we noticed inconsistencies between the timelines of ClinVar submissions and the cited publication dates, making it difficult to verify the most current and accurate evidence supporting reclassifications and compromising the trustworthiness of the evidence. Moreover, we observed patterns in cited studies, including bioinformatics tools and population diversity. These advancements are significantly enhancing the precision and efficiency of reclassification efforts.

## 2. Methods

### 2.1. Search Strategy

We investigated seven variants from a previous study [4], in which VUSs were reclassified as pathogenic or likely pathogenic between 2006 and 2016 (Figure 1). Following the American College of Medical Genetics and Genomics (ACMG) guidelines, which provide a standardized framework for classifying genomic variants from benign, likely benign, VUS, likely pathogenic, to pathogenic, each variant was searched on LitVar to obtain its dbSNP ID, cDNA nomenclature, and protein nomenclature. LitVar is a web service that employs advanced text mining and machine learning techniques to standardize variant names into unique IDs [5, 6]. We then searched for these standardized names in ClinVar to review variant submissions and their cited publications [3].

**Figure 1:**
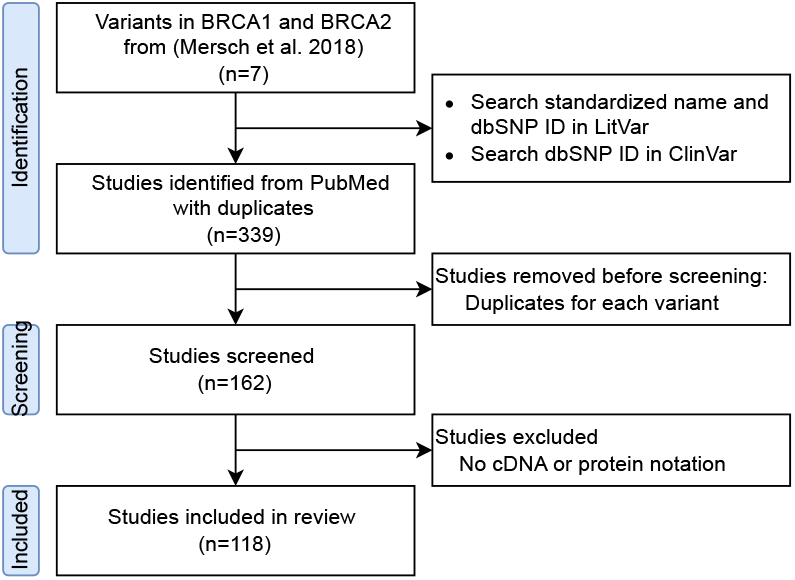
Study workflow.

### 2.2. Metadata Extraction

We extracted metadata such as publication year, population, and bioinformatics tools. Two annotators (HM and ZX) cross-verified this process, and a third one (YP) was consulted to resolve any disagreements.

## 3. Results and Discussion

### 3.1. Study Statistics

In this study, 162 articles were retrieved from ClinVar (Figure 1). Since a single article may report different variants, each was counted separately in our analysis. Consequently, we analyzed a total of 339 cited publications.

### 3.2. Unmatched ClinVar classification and cited publications

Table 1 shows the alignment between the variant classifications in ClinVar and those in the cited publications. Of the 339 cited publications, we found that 41 did not mention variants of interest in the main text, figures, tables, or supplements. 99 mentioned the variants, but not their classifications. Therefore, these 140 cited publications cannot be used to directly support all the criteria used for variant classifications in ClinVar, indicating a significant gap in reporting clarity and accuracy.

**Table 1.** Variant classifications submitted to ClinVar and their reported classifications in references.

| Classification in Clinvar | Classification in cited publications |  |  |  |  | Total (n=339) |
| --- | --- | --- | --- | --- | --- | --- |
|  | Pathogenic | Likely Path. | VUS | Unknown | Unfound |  |
| **Pathogenic** | 83 (30.3%) | 5 (1.8%) | 75 (27.4%) | 76 (27.7%) | 35 (12.8%) | 274 |
| **Likely pathogenic** | 11 (4.0%) | 1 (0.4%) | 19 (6.9%) | 21 (7.7%) | 3 (1.1%) | 55 |
| **VUS** | 2 (0.7%) | 2 (0.7%) | 1 (0.4%) | 2 (0.7%) | 3 (1.1%) | 10 |

Additionally, out of 274 cited publications used as evidence to support pathogenic variants, 5 actually reported the variants as Likely Pathogenic and 75 as VUS. More concerning, only one Likely Pathogenic instance was interpreted consistently in both ClinVar and in the cited article. In contrast, 11 publications were used to support Pathogenic and 19 for VUS classification in ClinVar. These findings reveal discrepancies between ClinVar and the literature, emphasizing the need for better alignment between ClinVar entries and the cited publications.

### 3.3. Inconsistent timelines between ClinVar and cited publications

We further compared the timelines of variant classification submissions in ClinVar with those reported in the literature (Figure 2). Here, we assume that in any year without submission or publication, the classification remains consistent with the previous year. The time range begins with the release of the first publication and ends with the release of the last publication. We observed that the timing of variant classification submissions in ClinVar often differs from what was reported in cited articles. For example, for variant c.5453A*>*G (Figure 2a), the “Pathogenic” classification appeared in ClinVar around 2010, yet it was not supported by any publication until 2018, indicating a five-year delay in evidence. Although we understand the community practice of submitting variant classifications based on unpublished data, it poses challenges for patients, clinicians, and researchers outside this field to comprehend the current evidence or build tools based on it, such as patient-facing chatbots. Similarly, Figure 2c shows that for variant c.4484G*>* A, the publication classified it as “Pathogenic” in 2010, while ClinVar did not reflect this classification until 2015, with fluctuations between the VUS and Pathogenic categories during these years.

**Figure 2.**
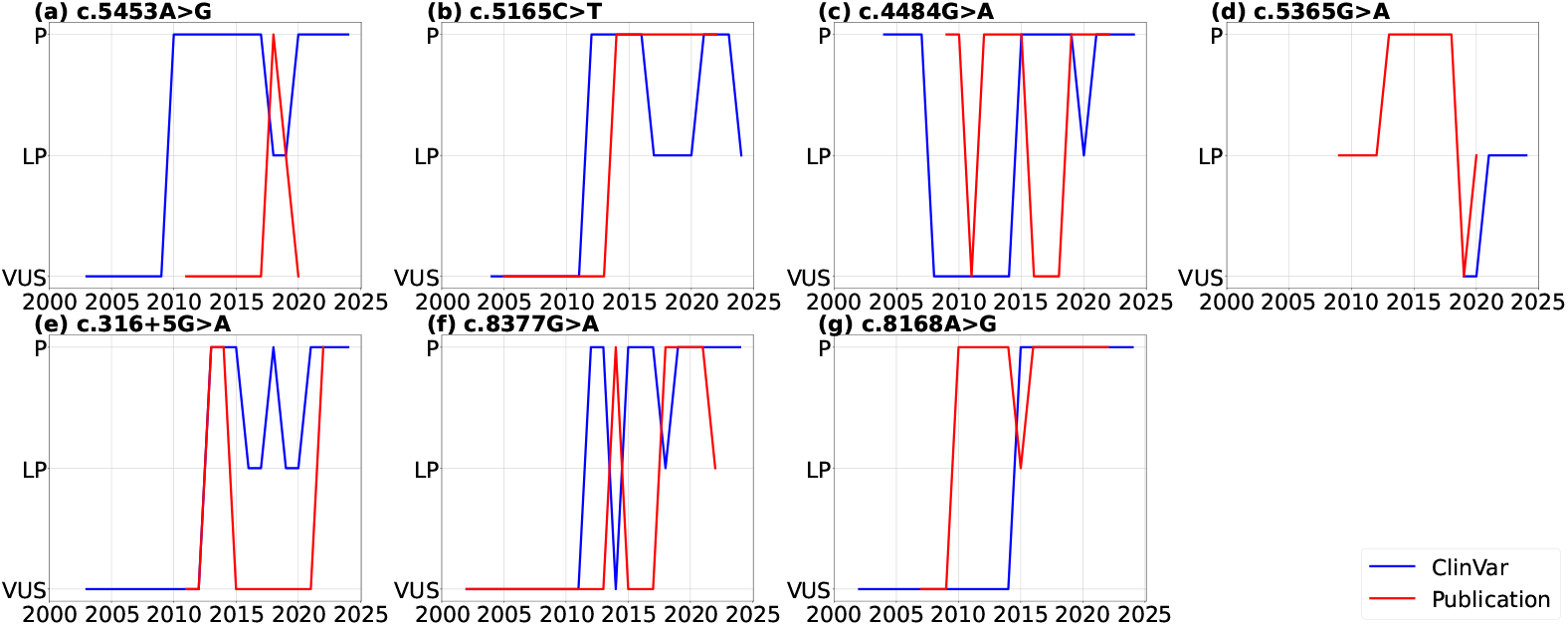
Comparison of variant classifications timelines on ClinVar with those reported in published literature (P: Pathogenic, LP: Likely Pathogenic, VUS: Variants of Uncertain Significance)

Potential reasons for these delays include reliance on unpublished data, resource constraints among submitters, and lag in manual curation or data integration. Such delays can hinder timely clinical decision-making, prolong uncertainty for patients, and complicate downstream efforts such as risk assessment, patient counseling, and tool development.

### 3.4. Observations on classification recommendations, genetic mechanism, and others

We first found that not all papers classified and interpreted variants following the recommendations of the American College of Medical Genetics (ACMG) and the International Agency for Research on Cancer (IARC) [7]. In some cases, research studies classified a variant as “deleterious” [8] or “likely deleterious” [9]. The discrepancy in terminology posed challenges in identifying consistent evidence in the included studies.

We then analyzed the factors that lead to variant reclassification. Biologically, among the seven variants we reviewed, all were missense variants or single amino acid substitutions. For example, c.8168A*>*G (p.Asp2723Gly) replaces aspartic acid with glycine at codon 2723 of the BRCA2 protein. Such missense variants can have clinical implications and often require the testing of additional family members and advanced modeling or functional validation. To assess whether the genetic variant disrupts gene function, computational analyses were carried out to assess the impact on mRNA splicing, which could predict potential deleterious effects on protein structure and function.

We identified five frequently used tools, including SIFT (Sorting Intolerant From Tolerant), NNSplice, Functional Assays, PolyPhen (Polymorphism Phenotyping), and MaxEntScan, which differ from the currently popular tools like CADD (Combined Annotation Dependent Depletion), REVEL (Rare Exome Variant Ensemble Learner), and alpha missense. A variety of tools across different studies are often used because a comprehensive test for pathogenicity usually requires multiple tools to ensure accurate classification. A combination of different tools increases confidence in predictions by accounting for multiple aspects of protein function. In some cases, functional assays provide definitive evidence to classify variants. However, when both computational and functional analysis demonstrated that a genetic variant has deleterious effects on the gene, it could be classified as pathogenic, according to the criteria of ACMG.

The publications also demonstrate the inclusion of diverse populations to explore the hereditary population aspects of breast and ovarian cancers. Participants were predominantly selected based on a strong family history of breast cancer. Key cohorts were sourced from regions such as India, France, Portugal, Denmark, Sweden, Italy, and the United States, with particular emphasis on ethnic subgroups, including Hispanic and Ashkenazi Jewish.

FAIR principles can facilitate variant reclassification by improving data findability, interoperability among tools, and reusability across populations. Adhering to these principles will help streamline evidence aggregation and promote consistency in classification efforts.

## 4. Conclusions

This meta-research study highlights conflicting and inconsistent interpretations of pathogenicity between ClinVar submissions and cited publications, as well as inconsistent publication timelines. These observations underscore the need for standardized criteria across sources. While technological advances in artificial intelligence (AI) and genomic sequencing offer promising opportunities to improve variant interpretation, addressing evidence gaps in population-specific data and functional validation remains a priority.

## Data Availability

All data produced are available online at ClinVar

## Acknowledgments

This project was supported by the National Library of Medicine under grant numbers R01LM014573 and R01LM014344, the National Human Genome Research Institute under grant number R01HG012655, and the National Center for Advancing Translational Sciences under grant number UL1TR002384.

